# Racial Differences in Liver Cancer Incidence and Risk Factors Among a Low Socioeconomic Population

**DOI:** 10.1101/2021.04.15.21255568

**Authors:** Sylvie Muhimpundu, Baqiyyah N. Conway, Shaneda Warren Andersen, Loren Lipworth, Mark D. Steinwandel, William J. Blot, Xiao-Ou Shu, Staci L. Sudenga

**Affiliations:** Division of Epidemiology, Vanderbilt University Medical Center, Nashville, TN, USA; School of Community and Rural Health, University of Texas Health Science Center at Tyler, Tyler, TX, USA; Department of Population Health Sciences, School of Medicine and Public Health, University of Wisconsin-Madison, Madison, WI, USA; University of Wisconsin Carbone Cancer Center, Madison, WI, USA; International Epidemiology Institute, Rockville, Maryland, USA

**Keywords:** cancer risk, southeast, racial disparity, SCCS

## Abstract

**Background:** Liver cancer incidence in the United States is higher among African Americans compared to Whites. The purpose of this study was to examine differences in risk factors associated with Hepatocellular carcinoma (HCC) among Whites and African Americans from low socioeconomic backgrounds in the Southern Community Cohort Study (SCCS).

**Methods:** The SCCS is a prospective cohort study with participants from the southeastern US. HCC incidence rates were calculated. Multivariable Cox regression was used to calculate HCC adjusted hazard ratios (aHR) associated with known baseline HCC risk factors for Whites and African Americans, separately.

**Results:** There were 294 incident HCC. The incidence rate ratio for HCCwas higher (IRR=1.4, 95%CI: 1.1-1.9) in African Americans compared to Whites. Whites saw a stronger association between self-reported Hepatitis C Virus (aHR= 19.24, 95%CI: 10.58-35.00) and diabetes (aHR= 3.55, 95%CI: 1.96-6.43) for the development of HCCcompared to African Americans (aHR= 7.73, 95%CI: 5.71-10.47 and aHR = 1.48, 95%CI: 1.06-2.06, respectively) even though the prevalence of these risk factors was similar between races. Smoking (aHR= 2.91, 95%CI: 1.87-4.52) and heavy alcohol consumption (aHR= 1.59, 95%CI: 1.19-2.11) were significantly associated with HCC risk among African Americans only.

**Conclusions:** In this large prospective cohort, we observed racial differences in HCC incidence and risk factors associated with HCC among African Americans and Whites.

**Impact:** Understanding HCC risk differences can assist prevention strategies that target people at high risk, potentially based on risk factors that differ by race.

## INTRODUCTION

The incidence of liver cancer has been shown to be increasing over the last decade in many parts of the world including the United States (US).(1-3) Liver cancer has a 5-year relative cancer survival rate at 18%, making it the fifth leading cause of cancer death in men and seventh in women in the US in 2016.(4, 5) By 2030, liver cancer is projected to be the third leading cause of cancer-related death in the US.(4) This increasing liver cancer incidence and mortality is likely due to the high prevalence of hepatitis C virus (HCV) infections in the “baby boomer” generation (born from 1945 to 1965 (6-8) and the increasing incidence of non-alcoholic fatty liver disease.(9)

There are differences in liver cancer incidence and mortality rates by race and ethnicity in the US. Liver cancer incidence rates are higher among African Americans (10.2 per 100,000) compared to Whites (6.3 per 100,000).(8) Although liver cancer stage at diagnosis is similar amongst African Americans and Whites, the 5-year survival in African Americans (21%) was significantly lower for all stages combined compared to Whites (25%).(8, 10) The determinants of racial disparities in liver cancer incidence are not clear. Observed differences in liver cancer trends may reflect race/ethnicity-specific differences in the prevalence of the risk factors for liver cancer. Known liver cancer risk factors include chronic HCV, chronic hepatitis B virus (HBV), nonalcoholic steatohepatitis, alcoholic liver disease, diabetes, and obesity.(8, 10-15) Hepatocellular carcinoma (HCC) accounts for 75-85% of liver cancers.(2) The purpose of this analysis is to examine racial differences in self-reported risk factors (e.g., smoking, alcohol, hepatitis infection, diabetes, and obesity) for HCC incidence among African Americans and Whites participating in the Southern Community Cohort Study (SCCS), a cohort of individuals with a low socioeconomic background.

## MATERIALS AND METHODS

### Study population

The SCCS is a prospective study designed to assess the determinants of racial and socioeconomic differences in cancer and other diseases.(16, 17) Participants aged 40 to 79 years were recruited across 12 states: Alabama, Arkansas, Florida, Georgia, Kentucky, Louisiana, Mississippi, North Carolina, South Carolina, Tennessee, Virginia, and West Virginia. Cohort recruitment took place between 2002-2009. Most participants (∼85%) were recruited from community health centers (CHCs) in the 12 states and completed a comprehensive baseline interview. The remainder of participants were recruited using stratified random samples of the residents in the 12 states and completed an identical mailed questionnaire. For this analysis, we chose to only include participants recruited from CHC to focus on persons that were underinsured or uninsured (Figure 1). The baseline survey collected self-reported information on diseases determinants like the participant’s demographic factors, history of prior medical conditions, diet and lifestyle. To be eligible, participants must have been English-speaking, and not under treatment for cancer within the past year (with the exception of non-melanoma skin cancer). The institutional review boards at Vanderbilt University Medical center and Meharry Medical College approved the study protocol.

**Figure 1.**
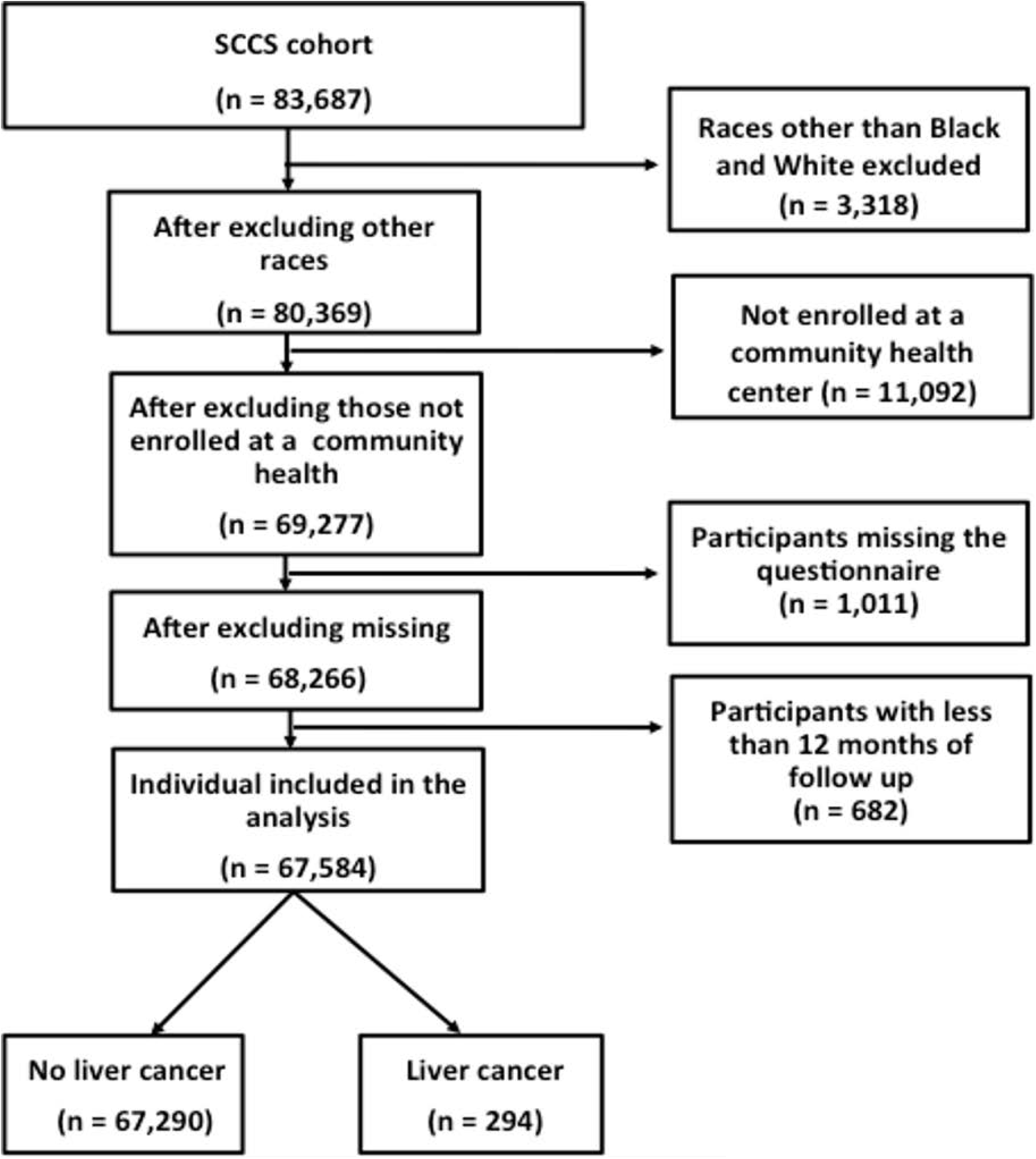
Flow chart of study participants. Exclusion and inclusion criteria for study participants is shown.

### Hepatocellular Carcinoma (HCC) Incidence

State Cancer Registries and the National Death Index (NDI) were the primary means of ascertainment of incident HCC diagnoses.(16) SCCS data can be linked with the 12 state cancer registries from which the participants were recruited. It is possible that participants can move out of those twelve states, so we also connect with NDI to identify those additional cases. Liver cancers were identified using the International Classification of Diseases (ICD) 10 code of C22.0 for hepatocellular carcinoma. The participants without HCC were linked to the Social Security Administration (SSA) for determination of current vital status. Cohort member information for those known through SSA to have died (or whose current status is unknown to SSA) was sent to the NDI to ascertain cause (or fact and cause) of death.(17) Cohort data was updated until December 31, 2017.

For participants with an incident HCC diagnosis, follow-up time was calculated from date of cohort enrollment to date of HCC diagnosis for those identified in cancer registry (n=270) or date of death from HCC for those identified in NDI (n=24). Follow-up time for participants without an incident HCC diagnosis was calculated from date of cohort enrollment to the date of death or date of vital status.

### Population for analysis

For the current study, we excluded participants with a prior history of liver cancer and those that were missing race and ethnicity (**Figure 1**). We also excluded participants not from Non-Hispanic White and Non-Hispanic African American ethnic groups (n=3,318), participants who were not enrolled at the community health centers (n=11,092), participants that did not have the baseline questionnaire (n= 1,011), or with a follow-up period of fewer than 12 months (n = 682). After all exclusions, 67,584 participants (72% Non-Hispanic African Americans and 28% Non-Hispanic Whites) were included in the analyses. We will refer to Non-Hispanic Whites as Whites and Non-Hispanic African Americans as African Americans for brevity. Participants self-identified as being African American or White. These self-identified racial categories were not based on genetic characterization. Baseline demographic information that was self-reported in the baseline survey was used to determine risk factors associated with HCC risk.

### Statistical analysis

We use baseline questionnaire responses for our exposures of interest. In this analysis, body mass index (BMI) was calculated as weight in kilograms (kg) over height (m^2^) and categorized as normal (<25 kg/m^2^), overweight (25 to 30 kg/m^2^) and obese (≥30 kg/m^2^). Self-reported age at baseline was analyzed using restricted cubic splines with 3 knots (White model) or 4 knots (African American model). Participants were asked how often they drank alcohol in the past year and how many drinks they typically had in a day when drinking. We categorized heavy alcohol drinkers as women with on average more than one alcoholic drink per day and men with on average more than two alcoholic drinks per day compared to women and men drinking less than that per day.(18) We analyzed alcohol both as a continuous variable and categorical variable and found similar results between the classifications. We report the analyses using categorical variable for alcohol in this manuscript. Current cigarette smoking status was classified as current, former, and never in our analyses. We assessed cigarette smoking intensity (pack-years) and found similar results to the cigarette smoking status. Hepatitis B virus (HBV), HCV and diabetes were all based on self-report with the participants being asked “Has a doctor ever told you that you have this virus/disease?” We did not ask if the participant had ever been tested for HBV, HCV or diabetes.

The age-adjusted incidence rate was calculated as the number of HCC incident cases divided by the follow up years, 95% confidence intervals (CIs) were estimated. Incidence rate ratios (IRR) and 95%CIs were also calculated comparing incidence rates between African Americans and Whites. Cox proportional hazards regression to compute hazard ratios (HRs) and 95% CIs for the incidence of HCC stratified by race. Known HCC risk factors and potential confounders (age, gender, BMI, education, household income, HBV, HCV, smoking, alcohol, employment status, and diabetes) were identified *a priori* and assessed within the models. We assessed the interaction between several known HCC risk factors, specifically between diabetes and HCV, and between smoking and alcohol. We also assessed the interaction between the known HCC risk factors and race. The proportionality assumption model using Schoenfeld residuals was assessed and held for the White model but did not hold for the African American model. When the African American model was stratified by gender, the proportionality assumption was satisfied. Separate unadjusted and adjusted models for African Americans by gender are provided as supplemental tables. All the data analysis was done using STATA/IC 15.0.

## RESULTS

### Study population

Overall, there were 294 incident HCC among Whites (n=57) and African Americans (n=237). Baseline characteristics of the study population by race are shown in **Table 1**. Whites and African Americans had similar prevalence of obese participants, 45% compared to 46%, respectively. Household income <$15,000 was more prominent in the African American population (62%) compared to the Whites (56%). Self-reported hepatitis C and hepatitis B infection prevalence was reported more often by Whites than African Americans. Diabetes prevalence was also similar between Whites (21%) and African Americans (22%). The prevalence of currently smoking was similar between African Americans (44%) and Whites (43%), but African Americans had a larger proportion that never smoked (36%) compared to Whites (31%). African Americans had a higher prevalence of heavy alcohol drinkers per day (20%) compared to Whites (11%). Being currently employed was higher among African Americans (38%) compared to Whites (33%). We found that ∼80% of both African American and Whites 65 and older were not currently employed.

**Table 1.** Baseline characteristics of the study population by race.

| Characteristics | White | African Americans |
| --- | --- | --- |
| Number at Risk | 18,678 | 48,906 |
| Number of liver cases | 57 | 237 |
| Follow up time (months; median (IQR)) | 110 (89-129) | 126 (100-145) |
| Age at enrollment (years; median (IQR)) | 52 (46-59) | 50 (44-56) |
| 40-44 | 20.1 | 25.3 |
| 45-49 | 21.5 | 24.5 |
| 50-54 | 18.5 | 20.7 |
| 55-59 | 15.0 | 13.0 |
| 60-64 | 11.7 | 8.02 |
| 65-69 | 7.0 | 4.6 |
| 70-74 | 4.0 | 2.7 |
| 75-79 | 2.3 | 1.4 |
| Sex, % |  |  |
| Male | 34.3 | 41.3 |
| Female | 65.7 | 58.7 |
| BMI category, % |  |  |
| Normal | 25.8 | 24.5 |
| Overweight | 28.6 | 28.8 |
| Obese | 44.7 | 45.7 |
| Missing | 0.8 | 1.1 |
| Education, % |  |  |
| < High School | 29.0 | 33.2 |
| High School | 35.4 | 35.2 |
| > High School | 35.6 | 31.6 |
| Missing | 0.0 | 0.0 |
| Household income, % |  |  |
| <\$15,000 | 56.0 | 61.8 |
| \$15,000-\$49,999 | 34.3 | 33.5 |
| >\$50,000 | 8.6 | 3.6 |
| Missing | 1.1 | 1.1 |
| Self-reported Hepatitis B Virus infection, % |  |  |
| No | 95.7 | 97.2 |
| Yes | 2.0 | 1.5 |
| Missing | 2.4 | 1.3 |
| Self-reported Hepatitis C virus infection, % |  |  |
| No | 92.6 | 95.6 |
| Yes | 5.0 | 3.1 |
| Missing | 2.4 | 1.3 |
| Diabetes, % |  |  |
| No | 79.0 | 77.8 |
| Yes | 20.7 | 22.0 |
| Missing | 0.3 | 1.3 |
| Among those with diabetes, use of prescription diabetes medication, % |  |  |
| No | 19.4 | 13.3 |
| Yes | 80.6 | 86.6 |
| Missing | 0.0 | 0.01 |
| Among those with diabetes, number of years with Diabetes (years, median (IQR)) | 6 (2-12) | 7 (2-14) |
| HIV, % |  |  |
| *No* | 98.9 | 45.1 |
| *Yes* | 0.8 | 2.0 |
| *Missing* | 0.3 | 0.2 |
| Insurance coverage, % |  |  |
| *No* | 43.3 | 43.2 |
| *Yes* | 56.0 | 56.3 |
| *Missing* | 0.7 | 0.6 |
| Use of Aspirin, % <sup>a</sup> |  |  |
| *No* | 85.3 | 88.2 |
| *Yes* | 14.3 | 11.5 |
| *Missing* | 0.4 | 0.4 |
| Use of over the counter pain-relievers, <sup>a</sup> % |  |  |
| *No* | 78.0 | 84.0 |
| *Yes* | 21.6 | 15.6 |
| *Missing* | 0.4 | 0.4 |
| Use of prescription pain-relievers, <sup>a</sup> % |  |  |
| *No* | 93.5 | 96.6 |
| *Yes* | 6.1 | 3.1 |
| *Missing* | 0.4 | 0.3 |
| Cigarette smoking status, % |  |  |
| *Never* | 30.8 | 36.4 |
| *Former* | 25.6 | 19.2 |
| *Current* | 43.4 | 44.2 |
| *Missing* | 0.2 | 0.2 |
| Heavy Alcohol drinkers, <sup>b</sup> % |  |  |
| *No* | 87.1 | 78.6 |
| *Yes* | 11.7 | 20.4 |
| *Missing* | 1.2 | 1.0 |
| Currently employed, % |  |  |
| *No* | 66.8 | 61.9 |
| *Yes* | 32.5 | 37.5 |
| *Missing* | 0.8 | 0.7 |
| Laborer, including construction worker, <sup>c</sup> % |  |  |
| *No* | 88.9 | 87.5 |
| *Yes* | 10.4 | 11.8 |
| *Missing* | 0.8 | 0.7 |
| Dry cleaning, <sup>d</sup> % |  |  |
| *No* | 98.7 | 98.3 |
| *Yes* | 0.6 | 1.0 |
| *Missing* | 0.8 | 0.7 |
| Farming, <sup>d</sup> % |  |  |
| *No* | 93.3 | 94.8 |
| *Yes* | 5.9 | 4.6 |
| *Missing* | 0.8 | 0.6 |
| Chemical production or use, <sup>d</sup> % |  |  |
| *No* | 96.01 | 97.2 |
| *Yes* | 3.2 | 2.1 |
| *Missing* | 0.8 | 0.7 |
| Abbreviations: IQR, Interquartile range; CHC, Community health clinic; BMI, Body Mass Index; Prescription pain relievers include Celebrex, Vioxx, or Bextra; HIV, Human Immunodeficiency Virus.<br><sup>a</sup> In the past year, participant has the person take the medication at least two times per week; <sup>b</sup> Heavy alcohol drinkers were defined as more than one drink on average for females and more than two drinks on average for males; <sup>c</sup> Laborer, including construction worker: whether they worked as laborers for the longest period of their adult life; <sup>d</sup> Farming, Dry cleaning, Chemical production or use: whether they worked in the corresponding field for more than 10 years. |  |  |

### Incidence rates

The overall age-adjusted incidence rate of HCC was 45.5 per 100,000 person-years (95%CI: 40.3-50.7) among our participants aged 40-79 living in the Southeastern United States. African Americans had higher age-adjusted incidence rates of HCC (IR=49.8 per 100,000 person-years) compared to Whites (IR=33.5 per 100,000 person-years) with an age-adjusted incidence rate ratio (IRR) of 1.48 (95%CI 1.1-2.0).

### Cox regression model

**Tables 2** and **3** show the multivariable-adjusted hazard ratios of HCC by race. After adjusting for potential risk factors and confounders with age included using restricted cubic splines, males were more likely to develop HCC than females in both African Americans (adjusted hazard ratio (aHR)= 2.05, 95%CI: 1.50-2.80) and Whites (aHR= 2.83, 95%CI: 1.58-5.07). HCV was significantly associated with HCC in both racial groups, with a more pronounced association among Whites (aHR= 19.24, 95%CI: 10.58-35.00) than African Americans (aHR= 7.73, 95%CI: 5.71-10.47). In combined race model assessing race and HCV and HCC, the interaction between race and HCV was not significant (p=0.08). Similarly, diabetes was significantly associated with HCC in both racial groups, but with a stronger association among Whites (aHR= 3.55, 95%CI: 1.96-6.43) than African Americans (aHR= 1.48, 95%CI: 1.06-2.06). The interaction between race and diabetes in a combined race model assessing race and diabetes and HCC was significant (p=0.01). Among African Americans, current cigarette smokers (aHR= 2.91, 95%CI: 1.87-4.52) had an increased risk of HCC compared to never smokers, but smoking was not significantly associated with HCC risk among Whites. In combined race model assessing race and smoking and HCC, the interaction between current smokers and race was significant (p=0.04). Furthermore, heavy alcohol consumption was significantly associated with HCC risk in African Americans only (aHR= 1.59, 95%CI: 1.19-2.11). BMI was not significantly associated with HCC in Whites and obese African Americans were at a lower risk for HCC (aHR= 0.65, 95%CI: 0.45-0.95).

**Table 2.**
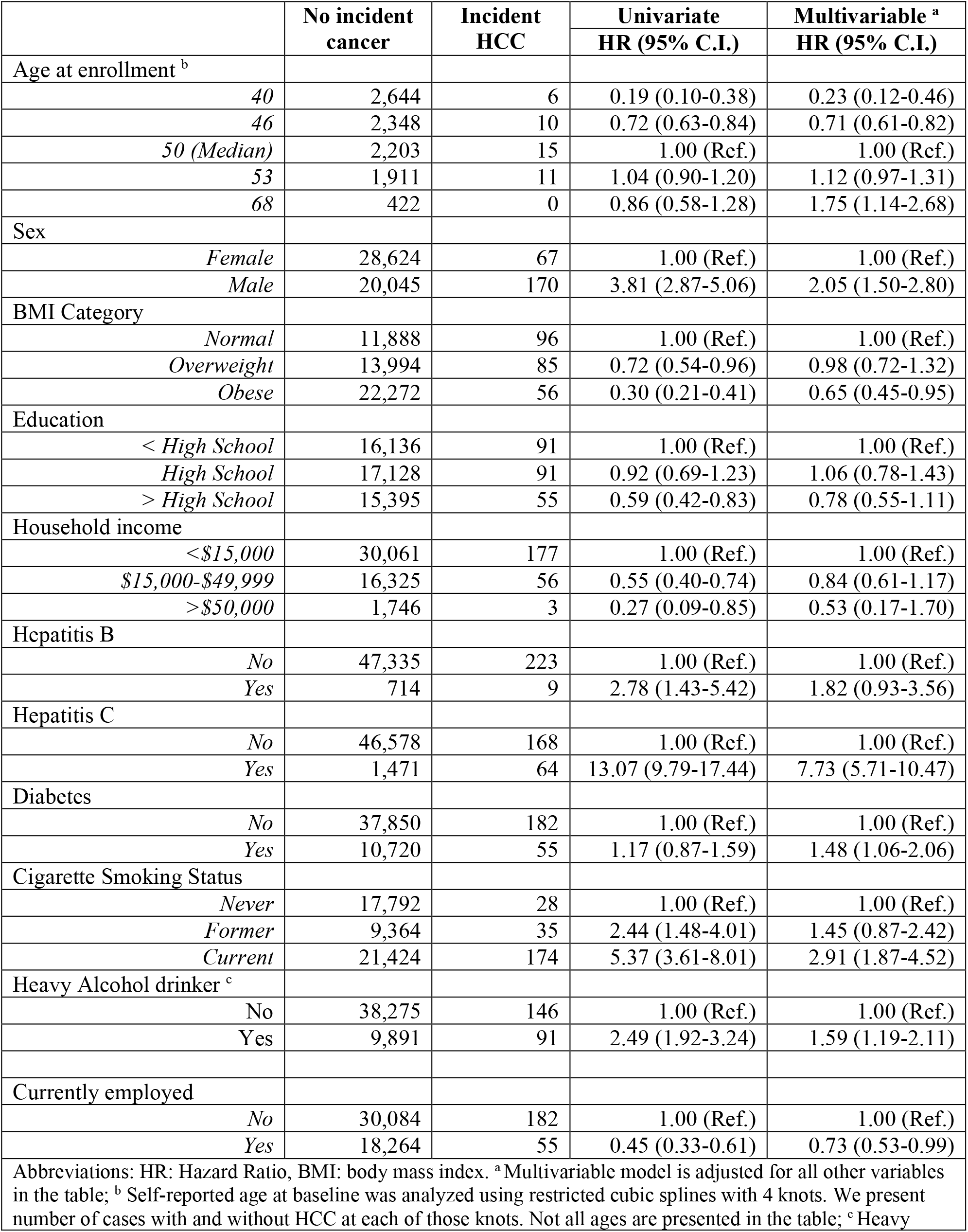
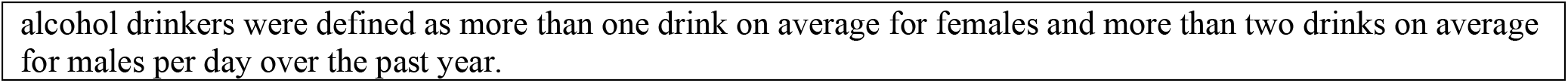
Univariate and multivariable hazard ratios (HR) and 95% confidence intervals (95% C.I.) for risk factors associated with Hepatocellular carcinoma (HCC) incidence among African Americans

|  | No incident cancer | Incident HCC | Univariate<br>HR (95% C.I.) | Multivariable <sup>a</sup><br>HR (95% C.I.) |
| --- | --- | --- | --- | --- |
| Age at enrollment <sup>b</sup> |  |  |  |  |
| 40 | 2,644 | 6 | 0.19 (0.10-0.38) | 0.23 (0.12-0.46) |
| 46 | 2,348 | 10 | 0.72 (0.63-0.84) | 0.71 (0.61-0.82) |
| 50 (Median) | 2,203 | 15 | 1.00 (Ref.) | 1.00 (Ref.) |
| 53 | 1,911 | 11 | 1.04 (0.90-1.20) | 1.12 (0.97-1.31) |
| 68 | 422 | 0 | 0.86 (0.58-1.28) | 1.75 (1.14-2.68) |
| Sex |  |  |  |  |
| Female | 28,624 | 67 | 1.00 (Ref.) | 1.00 (Ref.) |
| Male | 20,045 | 170 | 3.81 (2.87-5.06) | 2.05 (1.50-2.80) |
| BMI Category |  |  |  |  |
| Normal | 11,888 | 96 | 1.00 (Ref.) | 1.00 (Ref.) |
| Overweight | 13,994 | 85 | 0.72 (0.54-0.96) | 0.98 (0.72-1.32) |
| Obese | 22,272 | 56 | 0.30 (0.21-0.41) | 0.65 (0.45-0.95) |
| Education |  |  |  |  |
| < High School | 16,136 | 91 | 1.00 (Ref.) | 1.00 (Ref.) |
| High School | 17,128 | 91 | 0.92 (0.69-1.23) | 1.06 (0.78-1.43) |
| > High School | 15,395 | 55 | 0.59 (0.42-0.83) | 0.78 (0.55-1.11) |
| Household income |  |  |  |  |
| <\$15,000 | 30,061 | 177 | 1.00 (Ref.) | 1.00 (Ref.) |
| \$15,000-\$49,999 | 16,325 | 56 | 0.55 (0.40-0.74) | 0.84 (0.61-1.17) |
| >\$50,000 | 1,746 | 3 | 0.27 (0.09-0.85) | 0.53 (0.17-1.70) |
| Hepatitis B |  |  |  |  |
| No | 47,335 | 223 | 1.00 (Ref.) | 1.00 (Ref.) |
| Yes | 714 | 9 | 2.78 (1.43-5.42) | 1.82 (0.93-3.56) |
| Hepatitis C |  |  |  |  |
| No | 46,578 | 168 | 1.00 (Ref.) | 1.00 (Ref.) |
| Yes | 1,471 | 64 | 13.07 (9.79-17.44) | 7.73 (5.71-10.47) |
| Diabetes |  |  |  |  |
| No | 37,850 | 182 | 1.00 (Ref.) | 1.00 (Ref.) |
| Yes | 10,720 | 55 | 1.17 (0.87-1.59) | 1.48 (1.06-2.06) |
| Cigarette Smoking Status |  |  |  |  |
| Never | 17,792 | 28 | 1.00 (Ref.) | 1.00 (Ref.) |
| Former | 9,364 | 35 | 2.44 (1.48-4.01) | 1.45 (0.87-2.42) |
| Current | 21,424 | 174 | 5.37 (3.61-8.01) | 2.91 (1.87-4.52) |
| Heavy Alcohol drinker <sup>c</sup> |  |  |  |  |
| No | 38,275 | 146 | 1.00 (Ref.) | 1.00 (Ref.) |
| Yes | 9,891 | 91 | 2.49 (1.92-3.24) | 1.59 (1.19-2.11) |
| Currently employed |  |  |  |  |
| No | 30,084 | 182 | 1.00 (Ref.) | 1.00 (Ref.) |
| Yes | 18,264 | 55 | 0.45 (0.33-0.61) | 0.73 (0.53-0.99) |

Abbreviations: HR: Hazard Ratio, BMI: body mass index. <sup>a</sup> Multivariable model is adjusted for all other variables in the table; <sup>b</sup> Self-reported age at baseline was analyzed using restricted cubic splines with 4 knots. We present number of cases with and without HCC at each of those knots. Not all ages are presented in the table; <sup>c</sup> Heavy
alcohol drinkers were defined as more than one drink on average for females and more than two drinks on average for males per day over the past year.

**Table 3.** Univariate and multivariable hazard ratios (HR) and 95% confidence intervals (95% C.I.) for risk factors associated with Hepatocellular carcinoma (HCC) incidence among Whites

|  | No incident cancer | Incident HCC | Univariate<br>HR (95% C.I.) | Multivariable <sup>a</sup><br>HR (95% C.I.) |
| --- | --- | --- | --- | --- |
| Age at enrollment <sup>b</sup> |  |  |  |  |
| 42 | 763 | 0 | 0.52 (0.26-1.05) | 0.50 (0.24-1.04) |
| 52 (*Median*) | 665 | 6 | 1.00 (Ref.) | 1.00 (Ref.) |
| 66 | 313 | 1 | 0.64 (0.32-1.30) | 1.44 (0.69-3.01) |
| Sex |  |  |  |  |
| *Female* | 12,248 | 19 | 1.00 (Ref.) | 1.00 (Ref.) |
| *Male* | 6,373 | 38 | 4.24 (2.44-7.35) | 2.83 (1.58-5.07) |
| BMI Category |  |  |  |  |
| *Normal* | 4,806 | 21 | 1.00 (Ref.) | 1.00 (Ref.) |
| *Overweight* | 5,336 | 12 | 0.51 (0.25-1.03) | 0.52 (0.25-1.07) |
| *Obese* | 8,325 | 24 | 0.65 (0.36-1.16) | 0.79 (0.41-1.53) |
| Education |  |  |  |  |
| < *High School* | 5,395 | 20 | 1.00 (Ref.) | 1.00 (Ref.) |
| *High School* | 6,593 | 16 | 0.66 (0.34-1.27) | 0.67 (0.35-1.31) |
| > *High School* | 6,632 | 21 | 0.87 (0.47-0) | 0.82 (0.42-1.59) |
| Household income |  |  |  |  |
| < \$15,000 | 10,423 | 40 | 1.00 (Ref.) | 1.00 (Ref.) |
| \$15,000-\$49,999 | 6,393 | 10 | 0.39 (0.20-0.79) | 0.54 (0.26-1.12) |
| > \$50,000 | 1,601 | 7 | 1.14 (0.51-2.54) | 1.88 (0.74-4.75) |
| Hepatitis B |  |  |  |  |
| *No* | 17,817 | 53 | 1.00 (Ref.) | 1.00 (Ref.) |
| *Yes* | 361 | 4 | 3.83 (1.39-10.58) | 1.66 (0.58-4.73) |
| Hepatitis C |  |  |  |  |
| *No* | 17,275 | 28 | 1.00 (Ref.) | 1.00 (Ref.) |
| *Yes* | 903 | 29 | 22.41 (13.33-37.67) | 19.24 (10.58-35.00) |
| Diabetes |  |  |  |  |
| *No* | 14,714 | 35 | 1.00 (Ref.) | 1.00 (Ref.) |
| *Yes* | 3,849 | 22 | 2.61 (1.53-4.45) | 3.55 (1.96-6.43) |
| Cigarette Smoking Status |  |  |  |  |
| *Never* | 5,739 | 11 | 1.00 (Ref.) | 1.00 (Ref.) |
| *Former* | 4,770 | 12 | 1.37 (0.60-3.10) | 0.87 (0.38-2.00) |
| *Current* | 8,069 | 34 | 2.35 (1.19-4.65) | 1.23 (0.56-2.71) |
| Heavy alcohol drinker <sup>c</sup> |  |  |  |  |
| *No* | 16,234 | 42 | 1.00 (Ref.) | 1.00 (Ref.) |
| *Yes* | 2,164 | 15 | 2.53 (1.40-4.56) | 1.80 (0.95-3.42) |
| Currently employed |  |  |  |  |
| *No* | 12,438 | 38 | 1.00 (Ref.) | 1.00 (Ref.) |
| *Yes* | 6,043 | 19 | 0.96 (0.55-1.67) | 1.68 (0.90-3.14) |

Abbreviations: HR: Hazard Ratio, BMI: body mass index.
<sup>a</sup> Multivariable model is adjusted for all other variables in the table; <sup>b</sup> Self-reported age at baseline was analyzed using restricted cubic splines with 3 knots. We present number of cases with and without HCC at each of those knots. Not all ages are presented in the table; <sup>c</sup> Heavy alcohol drinkers were defined as more than one drink on average for females and more than two drinks on average for males per day over the past year.

## DISCUSSION

In this large ongoing cohort study of mostly low socioeconomic African American and White participants living in the southeastern U.S., we found racial differences in self-reported risk factors associated with HCC incidence. While the prevalence of several known risk factors for HCC were generally similar between African Americans and Whites, HCC incidence rates were higher among African Americans compared to Whites. Whites saw a greater effect between self-reported HCV and diabetes for the development of HCC compared to African Americans while smoking and alcohol use were significantly associated with HCC risk among African Americans only.

We found that males had a higher incidence of HCC than females in both African Americans and Whites. Gender differences in HCC incidence may reflect the higher prevalence of HCC risk factors in males than females (Supplemental Table 3). An additional possible explanation for the observed gender disparity in HCC incidence is the higher estrogen levels in females compared to males. Studies suggest that estrogen reduces HCC by inhibiting the production of Interleukin-6 (IL-6), a multifunctional cytokine that may be causal or contributory to HCC.(19-21)

In our study, self-reported HCV infection at baseline was associated with a 19-fold increased HCC incidence in Whites and 8-fold increase in African Americans in adjusted models. The overall self-reported HCV prevalence among Whites was 5.0% (95% CI 4.7%-5.3%) and 3.1% (95% CI 3.0%-3.3%) among African Americans. However, among Whites that developed HCC 50.9% (95% CI 37.9%-63.9%) were infected with HCV at baseline compared to 27.0% (95% CI 21.4%-32.7%) among African Americans. Previous studies found that although HCV is approximately 3-fold more prevalent in African Americans than in Whites, it does not cause liver diseases as severe in African Americans as in Whites.(22-24) In one retrospective study to determine the course of HCV in African Americans, HCV patients had a lower mean total Hepatic Activity Index (HAI), periportal hepatocyte necrosis scores, serum iron levels, and liver fibrosis scores compared to Whites with HCV. The finding suggests that African American chronic HCV patients have milder liver necroinflammation and fibrosis than White patients with similar HCV duration.(22, 24) A unique feature of HCC is its close association with liver fibrosis and cirrhosis, with more than 80% of HCCs developing in fibrotic or cirrhotic livers, suggesting an important role of liver fibrosis and cirrhosis in the premalignant environment of the liver. Studies have shown that among those with HCV, African Americans had a lower risk of cirrhosis (aHR= 0.58, 95 %CI: 0.55-0.60) and HCC (aHR= 0.77, 95 %CI: 0.71-0.83) compared with Whites.(25-27)

Alternatively, despite the recruitment of both African Americans and Whites from community health centers settings, it is possible that fewer African American SCCS participants were tested for HCV compared to White SCCS participants as HCV was self-reported in our cohort. However, without testing SCCS participants for HCV, it is unknown what the actual serostatus is in this population. NHANES data reporting HCV prevalence ratios comparing African Americans to Whites by state show higher HCV prevalence among African Americans compared to Whites in each of the 12 states included in the SCCS except for Mississippi.(28) At the time of enrollment, guidelines for HCV testing (29) in the US included a 1-time screening in all adults born between 1945 to 1965 and 80% of the SCCS population would qualify for this screening. The SCCS did not assess HCV testing in this population and therefore cannot make conclusion without testing and diagnosis information. Additionally, we do not have HCV treatment history available on these participants.

Previous studies have shown that diabetes is associated with a two-to three-fold increase risk of HCC and other conditions like chronic nonalcoholic liver disease.(30) These findings corroborate with our results; diabetes had a positive association with HCC. The association was statistically significant in both Whites and African Americans but was stronger in Whites compared to African Americans. Although the prevalence of self-reported diabetes at baseline was higher in African Americans 22.0% (95% CI 21.6%-22.4%) compared to Whites 20.7% (95% CI 20.1%-21.3%), the prevalence of self-reported diabetes was greater in Whites (38.6%, 95% CI 25.9%-51.2%) with an incident HCC compared with their counterpart in African Americans (23.2%, 95% CI 17.8%-28.6%). The difference in diabetes association with HCC between Whites and African Americans might be explained by the higher proportion of Whites with both HCV and diabetes at baseline (14.0%) that developed HCC compared to African Americans (6.8%) with both HCV and diabetes that developed HCC. However, this interaction was not statistically significant in our models. A previous study has found that there is a synergism on HCC risk among diabetes, chronic HBV/HCV infection, and alcohol consumption; presumably accelerating the process of fibrosis and progression to cirrhosis. The odds for HCC in patients with diabetes decreases when the patients do not have HBV, HCV and alcoholic liver diseases.(31)

Our findings with respect to both self-reported hepatitis and diabetes indicate that neither of these risk factors account for the observed higher incidence of HCC among African Americans than Whites in the SCCS population. Indeed, both illnesses were stronger determinants of risk among Whites than African Americans, so that the elevated HCC risk among African Americans than Whites occurs predominantly among those not affected by HCV or diabetes. Determinants of this disparity remain to be quantified.

A number of studies have reported an association between BMI and an increase in HCC risks.(32-34) A meta-analysis found that compared to individuals with normal weight (BMI 18.5–24.9 kg m^−2^), those who were overweight (BMI 25–30 kg m^−2^) or obese (≥30 kg m^−2^) had a 17% and 89%, respectively, increased risk of HCC.(32) We found no association between BMI and HCC risk in Whites or overweight African Americans in adjusted models. Obese African Americans were at a lower risk for HCC than normal weight African Americans. In our stratified analyses by sex among African Americans, the association between BMI and HCC risk was not significant in males but was for females. Difference in results may be due to BMI not capturing the association between obesity and HCC.(32) In one study, visceral fat and not BMI was associated with HCC risk.(35) The Multiethnic Cohort Study also found no association between HCC risk and obesity among African Americans, but did find an association among White males.(34) The study suggested that the previously reported relationship between BMI and the risk of HCC was mediated by visceral fat accumulation which is directly associated with hepatocarcinogenesis rather than BMI.(35) African Americans have been shown to have a lower amount of visceral fat compared to Whites.(36) Visceral abdominal adiposity was also associated with increased risk for hepatic steatosis and fibrosis in HCV as well as non-alcoholic fatty liver disease.(35)

Alcohol (37, 38) and cigarette smoking (11, 39) are well established risk factors for HCC. In this study, cigarette smoking and alcohol consumption were associated with HCC among African Americans but not Whites in adjusted models, although the interaction term between smoking and alcohol was not significant for Whites or African Americans. Cigarette smoking status was similar between African Americans and Whites at baseline while alcohol consumption was higher among African Americans compared to Whites. HCCSmoking cessation and reduction in alcohol consumption could potentially reduce the occurrence of HCC in African Americans.

Our study has several strengths and limitations. The strength of this study includes its prospective design, the use of a large data set from an ethnically diverse and low socioeconomic status cohort. Ascertainment of HCC was believed to be relatively complete via linkage with cancer registries in each of the 12 recruitment states and with the NDI. The study also considered most known and suspected HCC risk factors. Our results should also be read in the context of certain limitations. There were a small number of HCC cases among Whites which led to wide confidence intervals and there need to be caution in the interpretation of some of our findings. Several baseline characteristics were self-reported and subjected our analysis to recall errors. Hepatitis B and C were self-reported at baseline and there was no information on the treatment of these diseases. Our questionnaire did not capture information on liver diseases like cirrhosis, liver fibrosis, and nonalcoholic fatty liver disease and there could be racial differences in disease severity that could affect progression to HCC.

In conclusion, we have demonstrated that the prevalence and strength of the association with HCC for several known risk factors differed by race. The major risk factors of HCV, diabetes and obesity seem weaker (or in the case of obesity non-existent) among African Americans, suggesting that other factors account for the higher incidence rates of HCC among African Americans. It also is possible that other factors, such as genetic susceptibility, vary between African Americans and Whites and help explain the risk differentials. Further studies are needed to better understand the associations observed in this study and to identify factors that contribute to the racial differences in HCC risk.

## Data Availability

Data use approved by the Southern Community Cohort Study.

https://www.southerncommunitystudy.org/

## ACKNOWLEDGEMENT

Data on SCCS cancer cases used in this publication were provided by the Alabama Statewide Cancer Registry; Kentucky Cancer Registry, Lexington, KY; Tennessee Department of Health, Office of Cancer Surveillance; Florida Cancer Data System; North Carolina Central Cancer Registry, North Carolina Division of Public Health; Georgia Comprehensive Cancer Registry; Louisiana Tumor Registry; Mississippi Cancer Registry; South Carolina Central Cancer Registry; Virginia Department of Health, Virginia Cancer Registry; Arkansas Department of Health, Cancer Registry, 4815 W. Markham, Little Rock, AR 72205. The Arkansas Central Cancer Registry is fully funded by a grant from the National Program of Cancer Registries, Centers for Disease Control and Prevention (CDC). Data on SCCS cancer cases from Mississippi were collected by the Mississippi Cancer Registry, which participates in the National Program of Cancer Registries (NPCR) of the Centers for Disease Control and Prevention (CDC). Cancer data for SCCS cancer cases from West Virginia have been provided by the West Virginia Cancer Registry.

Research reported in this publication was supported by the National Cancer Institute of the National Institutes of Health. The content is solely the responsibility of the authors and does not necessarily represent the official views of the National Institutes of Health.

## References

1. Momin BR, Pinheiro PS, Carreira H, et al. Liver cancer survival in the United States by race and stage (2001-2009): Findings from the CONCORD-2 study. Cancer 2017;123 Suppl 24:5059–5078.

2. Bray F, Ferlay J, Soerjomataram I, et al. Global cancer statistics 2018: GLOBOCAN estimates of incidence and mortality worldwide for 36 cancers in 185 countries. CA Cancer J Clin 2018;68:394–424.

3. Torre LA, Siegel RL, Ward EM, et al. Global Cancer Incidence and Mortality Rates and Trends-- An Update. Cancer Epidemiol Biomarkers Prev 2016;25:16–27.

4. Rahib L, Smith BD, Aizenberg R, et al. Projecting cancer incidence and deaths to 2030: the unexpected burden of thyroid, liver, and pancreas cancers in the United States. Cancer Res 2014;74:2913–21.

5. Siegel RL, Miller KD, Jemal A. Cancer statistics, 2019. CA Cancer J Clin 2019;69:7–34.

6. Smith BD, Morgan RL, Beckett GA, et al. Recommendations for the identification of chronic hepatitis C virus infection among persons born during 1945-1965. MMWR Recomm Rep 2012;61:1–32.

7. Denniston MM, Jiles RB, Drobeniuc J, et al. Chronic hepatitis C virus infection in the United States, National Health and Nutrition Examination Survey 2003 to 2010. Ann Intern Med 2014;160:293–300.

8. Islami F, Miller KD, Siegel RL, et al. Disparities in liver cancer occurrence in the United States by race/ethnicity and state. CA Cancer J Clin 2017;67:273–289.

9. Kanwal F, Kramer JR, Mapakshi S, et al. Risk of Hepatocellular Cancer in Patients With Non-Alcoholic Fatty Liver Disease. Gastroenterology 2018;155:1828–1837.e2.

10. Ha J, Yan M, Aguilar M, et al. Race/ethnicity-specific disparities in cancer incidence, burden of disease, and overall survival among patients with hepatocellular carcinoma in the United States. Cancer 2016;122:2512–23.

11. Van Thiel DH, Ramadori G. Non-viral causes of hepatocellular carcinoma. J Gastrointest Cancer 2011;42:191–4.

12. Liu Y, Wu F. Global burden of aflatoxin-induced hepatocellular carcinoma: a risk assessment. Environ Health Perspect 2010;118:818–24.

13. Mittal S, El-Serag HB. Epidemiology of hepatocellular carcinoma: consider the population. J Clin Gastroenterol 2013;47 Suppl:S2–6.

14. Sahasrabuddhe VV, Gunja MZ, Graubard BI, et al. Nonsteroidal anti-inflammatory drug use, chronic liver disease, and hepatocellular carcinoma. J Natl Cancer Inst 2012;104:1808–14.

15. Bosch FX, Ribes J, Diaz M, et al. Primary liver cancer: worldwide incidence and trends. Gastroenterology 2004;127:S5–s16.

16. Signorello LB, Hargreaves MK, Blot WJ. The Southern Community Cohort Study: investigating health disparities. J Health Care Poor Underserved 2010;21:26–37.

17. Signorello LB, Hargreaves MK, Steinwandel MD, et al. Southern community cohort study: establishing a cohort to investigate health disparities. J Natl Med Assoc 2005;97:972–9.

18. US Department of Health Human Services. US Department of Agriculture. 2015–2020 dietary guidelines for Americans. In: Washington, DC; 2015.

19. Naugler WE, Sakurai T, Kim S, et al. Gender disparity in liver cancer due to sex differences in MyD88-dependent IL-6 production. Science 2007;317:121–4.

20. Villa E. Role of estrogen in liver cancer. Womens Health (Lond) 2008;4:41–50.

21. Baldissera VD, Alves AF, Almeida S, et al. Hepatocellular carcinoma and estrogen receptors: Polymorphisms and isoforms relations and implications. Med Hypotheses 2016;86:67–70.

22. Crosse K, Umeadi OG, Anania FA, et al. Racial differences in liver inflammation and fibrosis related to chronic hepatitis C. Clin Gastroenterol Hepatol 2004;2:463–8.

23. Conjeevaram HS, Kleiner DE, Everhart JE, et al. Race, insulin resistance and hepatic steatosis in chronic hepatitis C. Hepatology 2007;45:80–7.

24. Di Bisceglie AM, Lyra AC, Schwartz M, et al. Hepatitis C-related hepatocellular carcinoma in the United States: influence of ethnic status. Am J Gastroenterol 2003;98:2060–3.

25. El-Serag HB, Kramer J, Duan Z, et al. Racial differences in the progression to cirrhosis and hepatocellular carcinoma in HCV-infected veterans. Am J Gastroenterol 2014;109:1427–35.

26. Perz JF, Armstrong GL, Farrington LA, et al. The contributions of hepatitis B virus and hepatitis C virus infections to cirrhosis and primary liver cancer worldwide. J Hepatol 2006;45:529–38.

27. Veldt BJ, Chen W, Heathcote EJ, et al. Increased risk of hepatocellular carcinoma among patients with hepatitis C cirrhosis and diabetes mellitus. Hepatology 2008;47:1856–62.

28. Hall EW, Rosenberg ES, Sullivan PS. Estimates of state-level chronic hepatitis C virus infection, stratified by race and sex, United States, 2010. BMC Infectious Diseases 2018;18:224.

29. Moyer VA. Screening for hepatitis C virus infection in adults: U.S. Preventive Services Task Force recommendation statement. Ann Intern Med 2013;159:349–57.

30. Davila JA, Morgan RO, Shaib Y, et al. Diabetes increases the risk of hepatocellular carcinoma in the United States: a population based case control study. Gut 2005;54:533–9.

31. Yuan JM, Govindarajan S, Arakawa K, et al. Synergism of alcohol, diabetes, and viral hepatitis on the risk of hepatocellular carcinoma in blacks and whites in the U.S. Cancer 2004;101:1009–17.

32. Larsson SC, Wolk A. Overweight, obesity and risk of liver cancer: a meta-analysis of cohort studies. Br J Cancer 2007;97:1005–8.

33. Calle EE, Rodriguez C, Walker-Thurmond K, et al. Overweight, obesity, and mortality from cancer in a prospectively studied cohort of U.S. adults. N Engl J Med 2003;348:1625–38.

34. Setiawan VW, Lim U, Lipworth L, et al. Sex and Ethnic Differences in the Association of Obesity With Risk of Hepatocellular Carcinoma. Clin Gastroenterol Hepatol 2016;14:309–16.

35. Ohki T, Tateishi R, Shiina S, et al. Visceral fat accumulation is an independent risk factor for hepatocellular carcinoma recurrence after curative treatment in patients with suspected NASH. Gut 2009;58:839–44.

36. Carroll JF, Chiapa AL, Rodriquez M, et al. Visceral fat, waist circumference, and BMI: impact of race/ethnicity. Obesity (Silver Spring) 2008;16:600–7.

37. LoConte NK, Brewster AM, Kaur JS, et al. Alcohol and Cancer: A Statement of the American Society of Clinical Oncology. J Clin Oncol 2018;36:83–93.

38. Bagnardi V, Rota M, Botteri E, et al. Alcohol consumption and site-specific cancer risk: a comprehensive dose-response meta-analysis. Br J Cancer 2015;112:580–93.

39. Lee YC, Cohet C, Yang YC, et al. Meta-analysis of epidemiologic studies on cigarette smoking and liver cancer. Int J Epidemiol 2009;38:1497–511.

